# Spatial and temporal relationship between focal and rotational drivers and their relationship to structural remodeling in patients with persistent AF

**DOI:** 10.1101/2023.09.01.23294966

**Authors:** Shohreh Honarbakhsh, Caroline Roney, Amy Wharmby, Caterina Vidal Horrach, Ross J. Hunter

**Affiliations:** The Barts Heart Centre, Barts Health NHS Trust, Electrophysiology department, London, United Kingdom; Queen Mary University of London, London, United Kingdom

**Author notes:** Disclosures: Dr Honarbakhsh is a British Heart Foundation Clinical Intermediate Fellow and receives a British Heart Foundation Fellowship Grant. Dr Honarbakhsh has received speaker’s fees from Abbott. Professor Hunter has received research grants, educational grants, and speaker’s fees from Biosense Webster, Medtronic and Abbott. Dr Honarbakhsh and Professor Hunter are shareholders in Rhythm AI Ltd. Dr Roney acknowledges a UKRI Future Leaders Fellowship (MR/W004720/1). Funding: British Heart Foundation Clinical Intermediate Fellowship FS/ICRF/22/26034. Corresponding Author: Dr Shohreh Honarbakhsh MRCP BSc PhD, Cardiology and Electrophysiology Consultant, Senior Clinical Lecturer at Queen Mary University of London, The Barts Heart Centre, Barts Health NHS trust, W. Smithfield, London, EC1A 7BE.

**Keywords:** Atrial fibrillation, Localized drivers, Catheter ablation, Mapping

## Abstract

**Background:** Focal and rotational activations have been demonstrated in AF, but their relationship to each other and to structural remodeling remains unclear.

**Methods:** Patients undergoing catheter ablation for persistent AF were included. All patients underwent pulmonary vein isolation. Unipolar signals were collected to identify focal and rotational drivers using a wavefront propagation algorithm. Aim was to assess the relationship of drivers to underlying low voltage zones (LVZs; <0.5mV) and to determine whether there was a temporal (≤500ms) and spatial (≤12mm) relationship between focal and rotational drivers.

**Results:** In 40 patients, 86 drivers were identified (57, 66.3% focal and 29, 33.7% rotational). Rotational drivers showed co-localized to LVZs (21/29, 72.4%) whilst focal drivers did not (11/57 in LVZ, 19.3%; p<0.001). The proportion of the left atrium (LA) occupied by LVZs predicted rotational driver occurrence (AUC 0.96, 95%CI 0.90-1.00; p<0.001). In patients with a relatively healthy atrium, where the atrium was made up of ≤15% LVZs, only focal drivers were identified. Eighteen of the 21 (85.7%) rotational drivers located in LVZs also showed a temporal and spatial relationship to a focal driver. The presence of a LVZ within 12 mm of the focal driver was a strong predictor for whether a paired rotational driver would also occur in that vicinity.

**Conclusions:** Rotational drivers are largely confined to areas of structural remodelling and have a clear spatial and temporal relationship with focal drivers suggesting they are dependent on them. These novel mechanistic observations outline a plausible model for patient specific mechanisms maintaining AF.

## INTRODUCTION

It has been proposed that localized drivers maintain atrial fibrillation (AF). Several definitions have been used for these localized drivers, but two potential forms of localized drivers have been identified to date, focal and rotational. Studies have suggested that focal drivers and rotational drivers are both important in maintaining AF (1). Although the sites of these drivers appear to be patient specific, it is not clear whether there is a direct relationship between the two driver types or how they interact with the underlying atrial substrate, and this remains a large gap in our understanding of AF.

Drivers with rotational activity have a predilection for low voltage zones (LVZs) (1–4). It has long been proposed that functional reentry in the form of rotors may be responsible for this activity, and although rotors might gravitate towards areas of scarring, they may occur in healthy tissue (5,6). As functional reentry or anatomical reentry around a fixed obstacle such as scar are often initiated by premature electrical stimulation, it is possible that focal activity plays a role in initiating or maintaining these rotational drivers.

We hypothesized that rotational drivers occur predominantly in areas of structural remodeling and are initiated and supported by nearby focal drivers. To investigate this, rotational and focal drivers were mapped in patients with persistent AF and we hypothesized that there would be a spatial and temporal relationship between rotational and focal drivers, whereby rotational drivers would occur immediately following focal drivers when there is structural remodeling nearby.

## METHODS

Patients undergoing catheter ablation for persistent AF (<24 months and no previous AF ablation) were prospectively included. Exclusion criteria included age <18 years or reversible cause of AF. Patients provided informed consent for their study involvement which was approved by the UK National Research Ethics Service (22/PR/0961). The study was prospectively registered on clinicaltrials.gov (NCT05633303). Procedures were performed under either conscious sedation or general anaesthetic as per the clinician and patient’s preference. All procedures were performed on uninterrupted anticoagulation therapy with heparin administration during the procedure to maintain an activated clotting time (ACT) of >300 seconds.

### Electrophysiological mapping

Ensite X (Abbott, Chicago, IL, USA) was used as the 3D mapping system. Left atrial (LA) anatomical maps were created using the HD-grid mapping catheter (Abbott, Chicago, IL, USA). All patients had a high-density omnipolar voltage (OV) map created using the HD-grid catheter. Points that were ≥5mm from the geometry surface were filtered as not being in contact with the myocardium, and points acquired were respiratory gated to optimize the accuracy of anatomical localization. A minimum of 4000 bipolar voltage points were collected per patient with the aim of ensuring adequate atrial coverage. The interpolation threshold was set to 5mm for surface color projection and points were collected aiming for complete LA coverage (i.e., with no area >5mm from a data point). A decapolar catheter (Boston Scientific, MA, USA) was positioned in the coronary sinus (CS). The Tactiflex ablation catheter (Abbott, Chicago, IL, USA) was used for ablation.

### Ablation approach

Following voltage map creation, all patients underwent pulmonary vein isolation (PVI) with bilateral wide area circumferential ablation using radiofrequency ablation. PVI was achieved with lesions placed 5-10 mm outside the veno-atrial junction aiming for isolation as ipsilateral PV pairs. The anterior border of the left PVs was ablated on the LA appendage ridge where possible, or on the appendage side of the ridge where this was unstable. Lesions were delivered on the venous side of the appendage ridge only where this was necessary to isolate PVs. No further ablation was performed beyond PVI unless the patient organized into an atrial tachycardia (AT) or had previously documented AT in addition to AF. If the patients remained in AF, DC cardioversion (DCCV) was performed to achieve SR.

### Omnipolar voltage

For OV assessment, signals were obtained from three non-colinear electrodes that make up a clique. These are used to calculate the BV in all directions over 360 degrees. The BV with the largest bipolar peak-to-peak voltage is then used to compute a local virtual bipolar signal that represents the OV. Following this, all peak-to-peak voltage points identified within a 1mm sphere were identified. The voltage point with the highest OT certainty (numeric value ranging from 0-1 indicating how certain the calculated activation direction is) and largest voltage amplitude is then used for the final OV.

OV was used overcome the underestimation of voltage in AF due to impact of wavefront directionality and fractionation (7,8).

### Unipolar recordings

Following PVI, all patients had 30-seconds unipolar recordings collected using the HD-grid catheter. Unipolar recordings were collected by referencing to Wilson Central Terminal (WCT). A minimum of 30, 30-seconds unipolar recordings were collected to ensure adequate LA coverage. Unipolar recordings were overlapped to allow effective wavefront tracking. With each unipolar recording xyz coordinates were obtained for the electrode location to allow pairing of the electrodes to their neighboring electrodes and allow electrode position on the geometry to be determined.

The study aim was to assess the relationship between focal and rotational drivers in the LA and their relationship to structural LA remodeling. The RA was not analyzed.

### Offline analysis

#### i) Identification of focal drivers and rotational drivers

A wavefront tracking algorithm developed and executed in Matlab (Mathworks, MA, USA) was used offline to identify focal and rotational drivers. Unipolar electrograms, electrode xyz coordinates and geometry data were utilized. Firstly, ventricular far field signals were filtered. Atrial signals were then annotated. Filtering of far field ventricular and atrial signals were evaluated manually to ensure accurate exclusion of far field signals and atrial signal annotation. Inaccurate atrial annotations due to noise or fractionated signals were excluded from the analysis.

Electrodes were then paired to their neighboring electrodes through comparing geodesic distance between electrodes. The atrial activations were then compared amongst these electrodes to track the wavefront propagation. Firstly, stable wavefront propagation was identified. A stable wavefront propagation was defined as one that demonstrated consecutive repetition ≥2 cycles with >2 repetitions over the 30-second recording. Combining multiple overlapping unipolar recordings allowed a prediction of the wavefront propagation over a segment to be determined. The stable wavefront propagations were then evaluated to identify a focal and rotational driver. For a focal driver to be identified the wavefront tracking algorithm needed to **i)** demonstrate a radial spread of wavefront i.e., wavefront spread from the electrode to neighboring electrodes at ≥120 degrees **ii)** the electrode demonstrating earlier atrial signal compared to neighboring electrodes demonstrating a QS morphology on the unipolar recordings. To identify a rotational driver **i)** a series of electrograms in consecutive electrodes needed to occupy more than 50% of the local cycle length **ii)** with less than 20mm between the starting and ending point of the activation. Two or more such activations occurring where at least 80% of the electrodes followed the same propagation pattern were defined as rotational activation (9). The methodology used is summarized in Figure 1.

**Figure 1.**
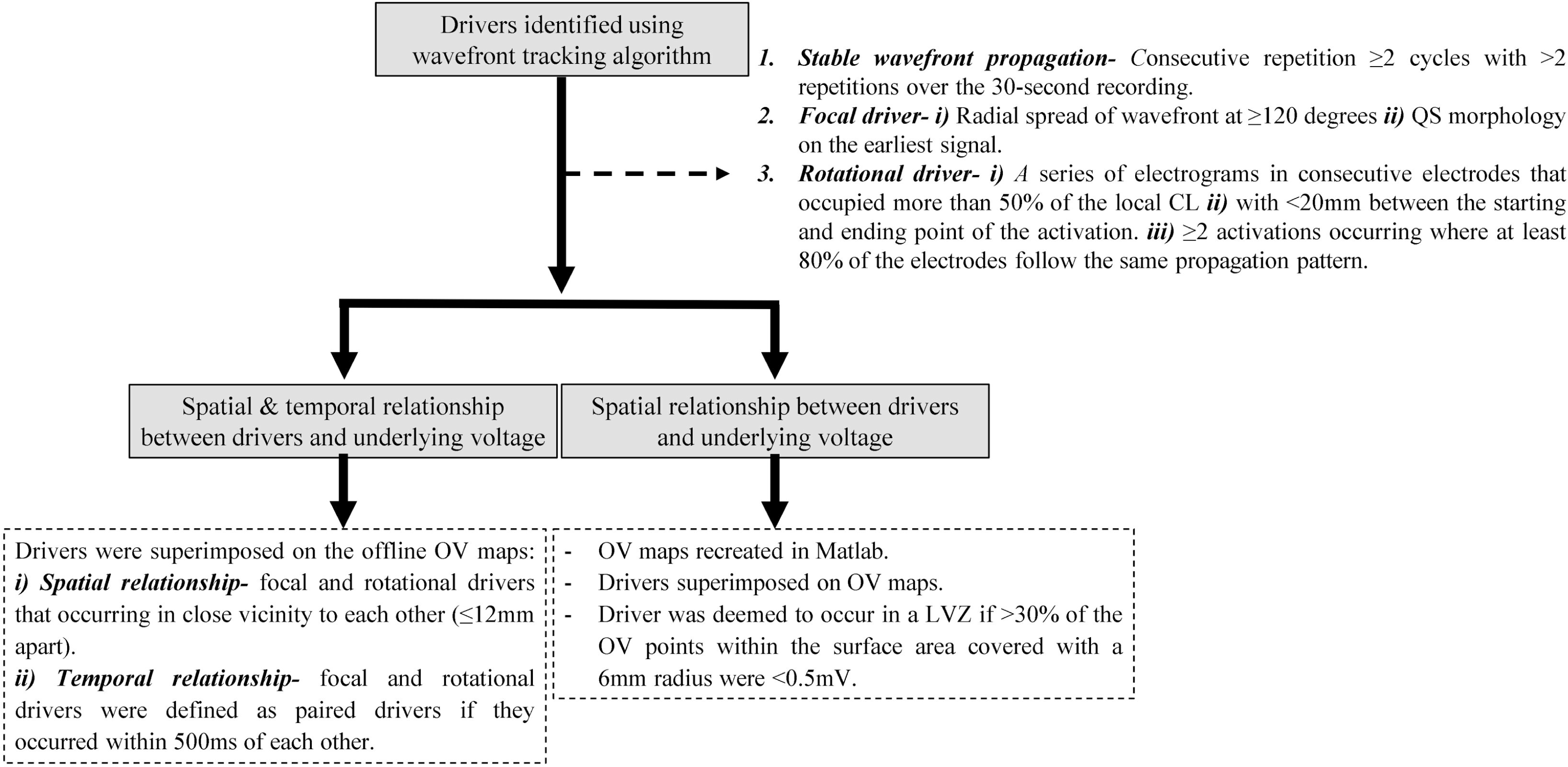
The flow diagram demonstrates the study methodology used.

#### ii) Anatomical distribution of drivers

The distribution of the drivers on a per anatomical segments were evaluated. A six-segment model was used sub-dividing the LA into lateral, septum, anterior, posterior, inferior and roof.

#### iii) Driver relationship with underlying omnipolar voltage

The OV maps were re-created in Matlab and the driver sites were superimposed on the OV maps for analysis of the spatial relationship between LVZs and the different driver types. The underlying OV was determined at the driver sites and was defined as either non-LVZs (nLVZs; ≥0.5 mV) or LVZs (<0.5 mV) (10). The voltage point was then projected onto a replica of the anatomy created in EnsiteX and each point was ascribed a surface area with a 5mm radius. Points with a voltage of <0.5mV were colored red whilst green was indicative of a voltage of ≥0.5mV. The driver was deemed to occur in a LVZ if >30% of the OV points within the surface area covered with a 6mm radius were <0.5mV.

Since rotational drivers may occupy a wider area than focal drivers, the relationship between rotational drivers and underlying structural remodeling was also determined through **i)** calculating the surface area covered by the rotational driver and then **ii)** determining the proportion of LVZs within this surface area. The surface area covered by the rotational driver was determined through tracing the inner border of the rotational activity. Within this designated area, all bipolar voltage points were identified and the proportion of these that were <0.5 mV was determined. The rotational driver was deemed to occur in a LVZ if >30% of the bipolar voltage points within the surface area were <0.5mV.

#### iv) Spatial relationship between focal and rotational drivers

Drivers were superimposed on the offline OV maps. For focal drivers the xyz coordinates of the leading electrode with a QS morphology was used to display the position on the OV maps. For rotational drivers the center of the traced rotational driver was used to display the position on the OV maps. Focal and rotational drivers were said to have a spatial relationship and were termed paired if they occurred within a 12mm geodesic distance of one another measured center to center. A geodesic distance of 12mm was used by considering the electrode distance between the center of a HD grid catheter to the center of another HD-grid catheter which measures between 6-12 mm. The maximum measurement was utilized. The underlying voltage at these sites was determined.

#### v) Temporal relationship between focal and rotational drivers

The temporal relationship between focal and rotational drivers was assessed by reviewing wavefront tracking algorithms and their corresponding electrograms. Focal and rotational drivers were defined as paired drivers if they occurred within 500ms of each other.

#### vi) Driver characteristics

For each driver the temporal stability and recurrence rate was determined using the wavefront propagation algorithm. Temporal stability was defined as the mean number of consecutive repetitions identified during each occurrence of a driver. The number of times a driver was identified and met the study definition of driver during a 30-second recording was defined as the recurrence rate (2,11).

### Statistical analyses

Statistical analyses were performed using SPSS (IBM SPSS Statistics, Version 25 IBM Corp, NY, USA). Continuous variables are displayed as mean ± standard deviation (SD) or median (range). Categorical variables are presented as numbers and percentages. The Student T-test or Mann-Whitney U test was used for comparison of continuous variables. Spearman rank correlation coefficient was determined to assess the relationship between AF duration and proportion of LVZs. Binary logistic regression was used to identify predictors of rotational drivers. Area under the curve (AUC) was calculated to determine if the proportion of LVZs could predict the driver type identified during mapping. P-value of <0.05 was deemed significant.

## RESULTS

The study included 40 patients with a mean age of 60.5±11.9 years and predominantly male (n=30, 75.0%). The mean AF duration was 17.1±6.7 months. Out of these patients, 14 (35.0%) had an AF duration ≤12 months and 26 (65.0%) had an AF duration >12 months. Baseline characteristics are demonstrated in Table 1. All patients were in AF at the start of their procedure. No complications were encountered. Post PVI all patients remained in AF and underwent DCCV to SR. No further ablation beyond PVI was performed.

**Table 1.**
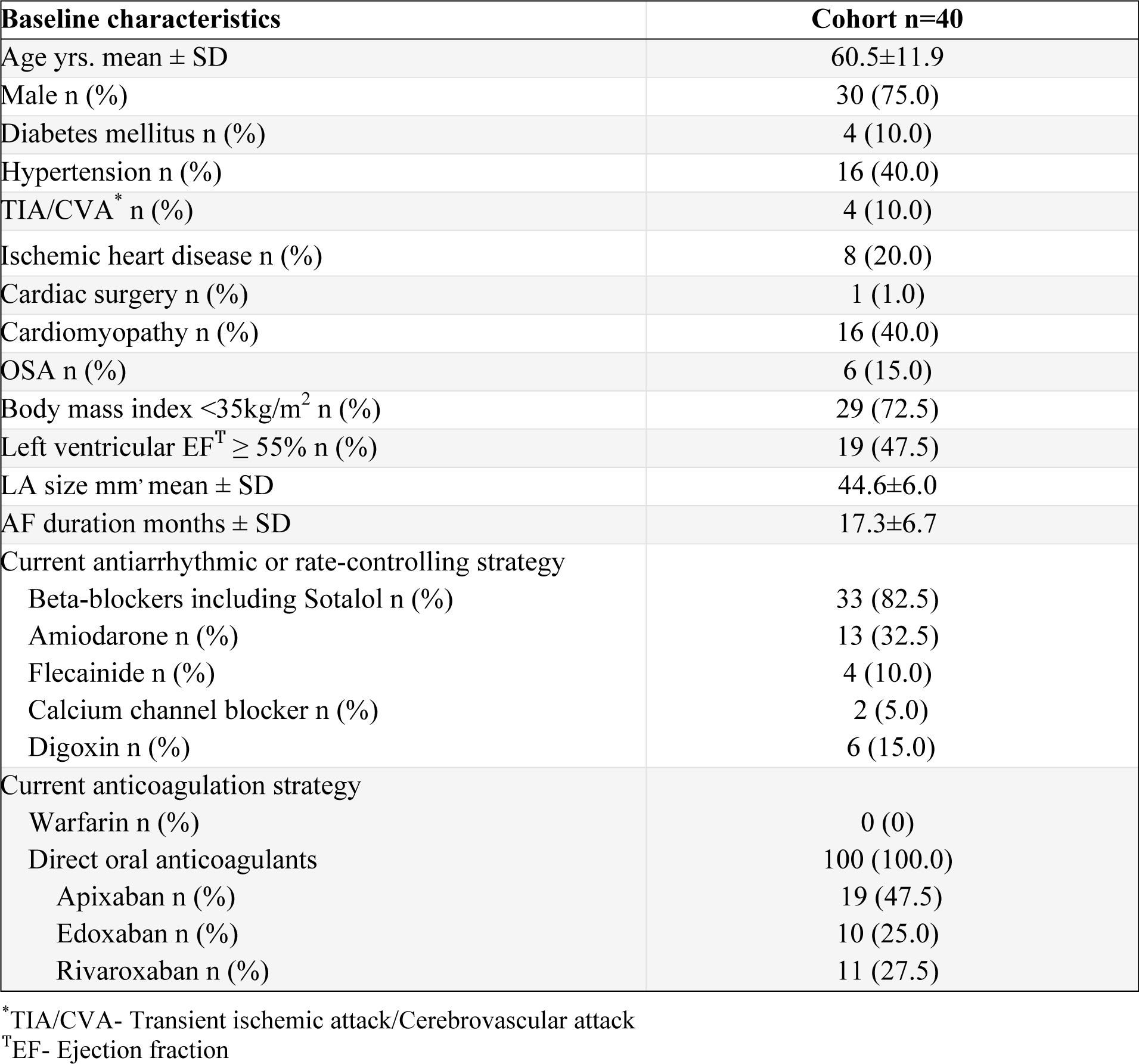
Baseline characteristics.

| Baseline characteristics | Cohort n=40 |
| --- | --- |
| Age yrs. mean ± SD | 60.5±11.9 |
| Male n (%) | 30 (75.0) |
| Diabetes mellitus n (%) | 4 (10.0) |
| Hypertension n (%) | 16 (40.0) |
| TIA/CVA* n (%) | 4 (10.0) |
| Ischemic heart disease n (%) | 8 (20.0) |
| Cardiac surgery n (%) | 1 (1.0) |
| Cardiomyopathy n (%) | 16 (40.0) |
| OSA n (%) | 6 (15.0) |
| Body mass index $<35\text{kg/m}^2$ n (%) | 29 (72.5) |
| Left ventricular EF<sup>T</sup> $\geq 55\%$ n (%) | 19 (47.5) |
| LA size mm' mean ± SD | 44.6±6.0 |
| AF duration months ± SD | 17.3±6.7 |
| Current antiarrhythmic or rate-controlling strategy |  |
| Beta-blockers including Sotalol n (%) | 33 (82.5) |
| Amiodarone n (%) | 13 (32.5) |
| Flecainide n (%) | 4 (10.0) |
| Calcium channel blocker n (%) | 2 (5.0) |
| Digoxin n (%) | 6 (15.0) |
| Current anticoagulation strategy |  |
| Warfarin n (%) | 0 (0) |
| Direct oral anticoagulants | 100 (100.0) |
| Apixaban n (%) | 19 (47.5) |
| Edoxaban n (%) | 10 (25.0) |
| Rivaroxaban n (%) | 11 (27.5) |

\*TIA/CVA- Transient ischemic attack/Cerebrovascular attack
<sup>T</sup>EF- Ejection fraction

**Table 2.** Demonstrates the characteristics of focal and rotational drivers identified.

|  | **Focal driver** | **Rotational driver** |
| --- | --- | --- |
| Driver type n % | 57 | 48 |
| LVZs<sup>*</sup> n % | 11 (19.3) | 13 (27.1) |
| nLVZs<sup>T</sup> n % | 46 (80.7) | 35 (72.9) |
| Temporal stability mean ± SD | 4.0±0.6 | 3.0±0.6 |
| Recurrence rate mean ± SD | 4.8±1.7 | 3.3±1.9 |
| Paired with another driver type | 32 (69.6) | 32 (91.4) |

\*LVZs- Low voltage zones
<sup>T</sup>nLVZs- Non low voltage zones

### Identification of localized drivers

In the 40 patients, 105 drivers were identified using the novel wavefront propagation maps (2.6±0.9 per patient). All patients had drivers identified. Out of the 105 localized drivers identified, 57 (66.3%) were focal in nature and 48 (45.7%) were rotational drivers.

### Characteristics of drivers

During a 30-second recording, drivers demonstrated a mean temporal stability of 3.6±0.8 consecutive repetitions. Focal drivers showed greater temporal stability than rotational drivers did (4.2±0.5 vs. 2.9±0.8; p<0.001). During a 30-second recording, the drivers showed on average a recurrence rate of 8.7±3.2 times. Focal drivers demonstrated a higher recurrence rate than rotational drivers (12.1±3.1 vs. 7.1±4.6; p<0.001).

A majority of the drivers identified were mapped to the anterior wall (n=29; 27.6%), roof (n=23, 21.9), posterior wall (n=16; 15.2%), lateral wall (n=14, 13.3%), septum (n=12, 11.4%) and inferior wall (n=11, 10.5%). The anatomical distribution was consistent between both focal and rotational drivers (Figure 2A-B).

**Figure 2A-B.**
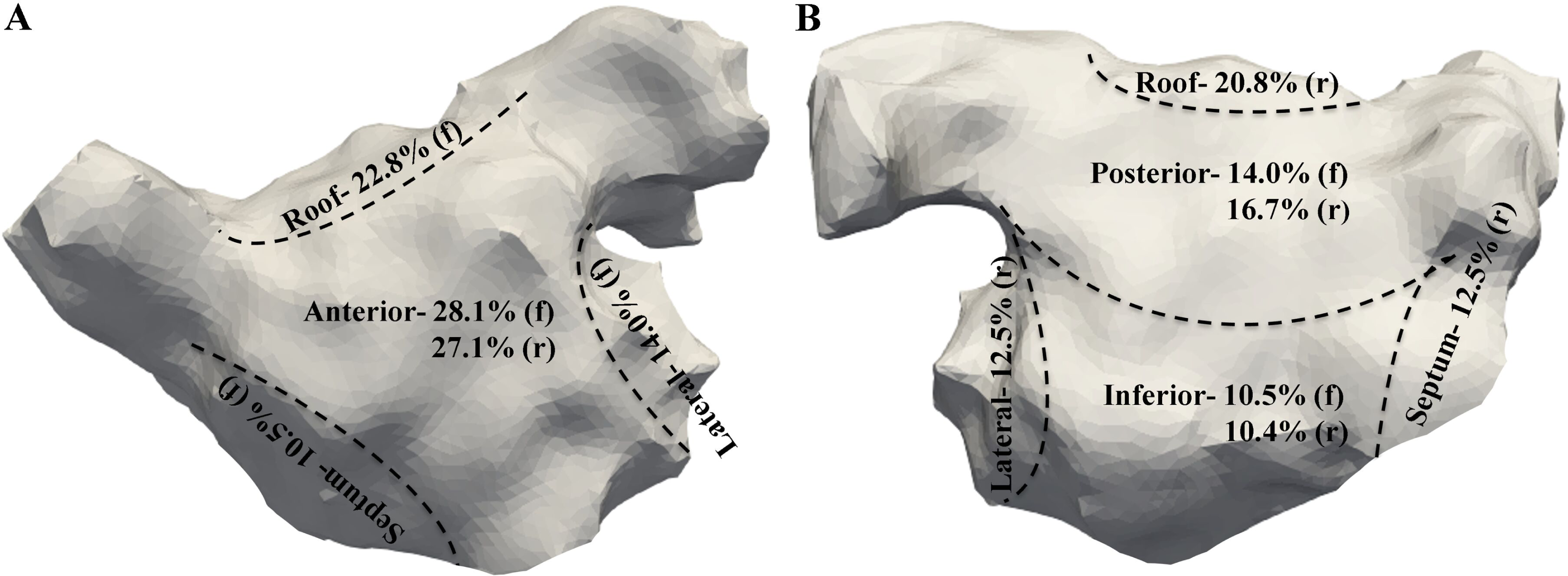
Demonstrates the anatomical distribution of ***A-*** Focal drivers (f) ***B-*** Rotational drivers (r) on a 6-segment model (anterior, roof, septum, lateral, posterior, and inferior wall).

### Underlying bipolar voltage zone at driver sites

OV maps in AF were available in all 40 patients. The average number of voltage points per patient were 77219.6±51075.7 points. The proportion of LA surface area consisting of LVZs in those with an AF duration of <12 months was 16.4±4.5% whilst in those with an AF duration of >12months it was 30.1±7.8%; p<0.001. There was a strong positive correlation between AF duration and proportion of LVZs (r_s_=0.93; p<0.001).

#### i) Focal drivers

In total, 37 patients (92.5%) had focal drivers identified (1.5±0.7 per patient). Out of the 57 focal drivers identified a majority were mapped to nLVZs (n=46, 80.7%). There was also no difference in temporal stability (4.1±0.5 vs. 3.9±0.7; p=0.78) or recurrence rate (4.7±1.9 vs. 4.9±1.6; p=0.65) of focal drivers mapped to LVZs vs. those mapped to nLVZs. Proportion of nLVZs was a significant predictor of identifying focal drivers (Odds ratio 1.15, 95%CI 1.04-1.26; p=0.005). The proportion of the LA occupied by nLVZs showed an AUC of 0.78 (95%CI 0.64-0.93; p=0.003) in predicting the presence of focal drivers with an optimal cutoff of 74.5% (Sensitivity 81.3% and Specificity 72.0%). In patients with a relatively healthy atrium, where the atrium was made up of ≤15% LVZs, only focal drivers were identified.

#### ii) Rotational drivers

In total, 30 patients (75.0%) had rotational drivers identified (1.6±0.3 per patient). Out of the 48 rotational drivers, 35 (72.9%) were mapped to LVZs (0.31±1.2mV). Whilst rotational drivers showed a predilection to LVZs, focal drivers did not (p<0.001). Proportion of LVZs was a significant predictor of identifying rotational drivers (Odds ratio 1.78, 95%CI 1.09-2.91; p=0.02). The proportion of the LA occupied by LVZs showed an AUC of 0.96 (95%CI 0.90-1.00; p<0.001) in predicting the presence of rotational driver with an optimal cutoff of 19.5% (Sensitivity 92.9% and Specificity 76.9%).

Rotational drivers mapped to LVZs demonstrated greater temporal stability (3.3±0.8 vs. 2.6±0.5, p=0.002) and recurrence rate (3.9±1.8 vs. 2.7±2.0; p=0.002) compared to rotational drivers mapped to nLVZs.

### Relationship between focal and rotational drivers

Focal and rotational drivers occurring in close proximity to each other (≤12mm) was seen for 32 pairs i.e., 64 out of the 105 (61.0%) drivers identified. Thirty-two out of 35 (91.4%) rotational drivers were mapped in close proximity to a focal driver whilst 32 out of the 57 (56.1%) focal drivers were mapped to an area in close proximity to a rotational driver (p<0.001) (Figure 3A-D and Figure 4A-B).

**Figure 3A-C.**
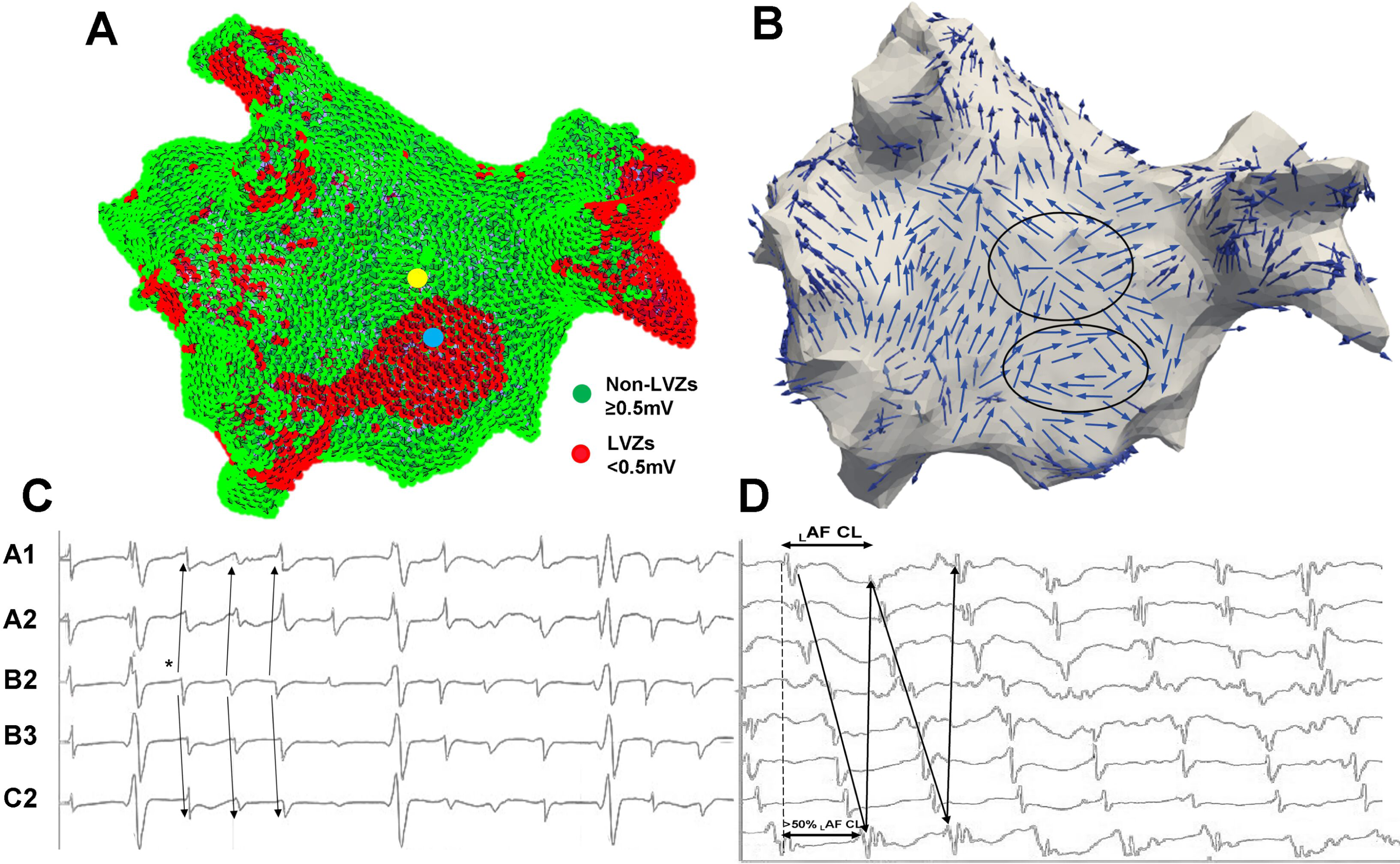
Demonstrates ***A-*** A replica geometry and OV map created in EnsiteX in a posterior-anterior view (green highlights nLVZ (≥0.5mV) and red highlights LVZs (<0.5mV)). A focal driver was mapped to a nLVZ (yellow circle), and a rotational driver was mapped in close proximity to a LVZ (blue circle). ***B-*** Wavefront propagation map in a posterior-anterior view created using the unipolar recordings. The top black circle highlights the wavefront propagation of the focal driver, and the lower black circle highlights the wavefront propagation of the rotational driver. ***C-*** Demonstrates the electrograms obtained at the focal driver site. This demonstrates a QS morphology at the leading electrode (*) with radial spread in activation to neighboring electrodes. ***D-*** Demonstrates the electrograms obtained at the rotational driver site. This demonstrates the spread of activation across consecutive electrodes occupying more than 50% of the local cycle length with more than 2 such activations.

**Figure 4A-B.**
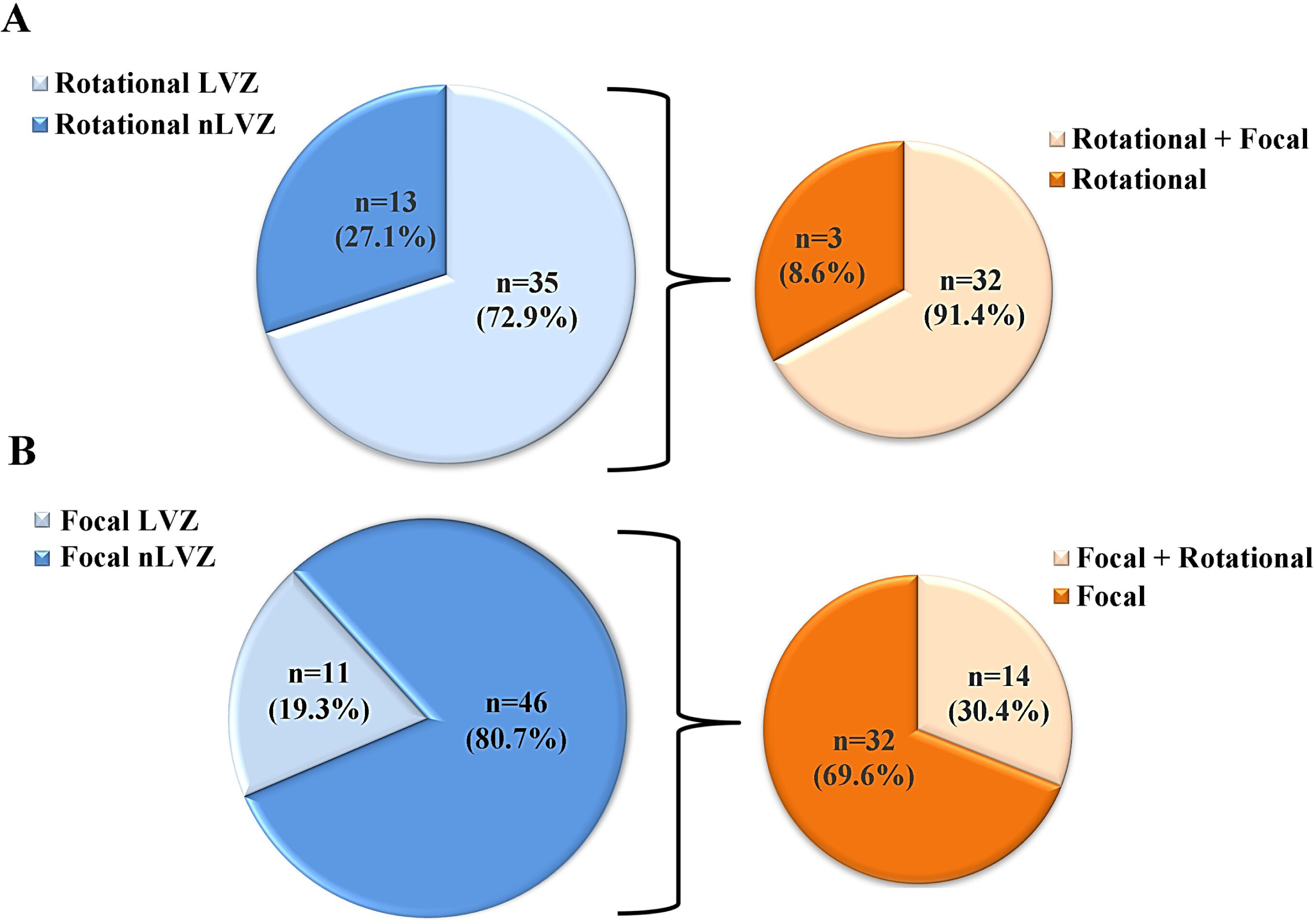
Demonstrates pie charts that summarize the distribution of ***Ai-*** Rotational drivers in accordance with those mapped to LVZ (light blue) and nLVZs (dark blue) and ***Aii-*** The proportion of the rotational drivers that occurred in close proximity to focal drivers (light orange) and the proportion that did not (dark orange). All rotational drivers that occurred in close proximity to focal drivers were mapped to LVZs. ***Bi-*** Focal drivers in accordance with those mapped to LVZ (light blue) and nLVZs (dark blue) and ***Bii-*** The proportion of the focal drivers that occurred in close proximity to rotational drivers (light orange) and the proportion that did not (dark orange). All focal drivers that occurred in close proximity to rotational drivers were mapped to nLVZs.

Beyond demonstrating a spatial relationship, focal and rotational drivers that occurred in close proximity to each other also demonstrated a temporal relationship. Out of the 32 pairs, 29 focal drivers occurred within 500ms before the occurrence of the rotational driver (90.6%) with the focal drivers occurring on average 389±65ms before the rotational driver measured from the onset of the focal driver. The rotational driver occurred on average following 2.5±0.7 cycles of the focal driver.

### Relationship between LVZs and the pairing of focal and rotational drivers

All the 32 paired focal drivers were mapped to a nLVZ whilst all of the 32 paired rotational drivers were mapped to a LVZ <12 mm away (Figure 5A-D). When considering all 57 focal drivers, the presence of a LVZ within 12 mm was a strong predictor for whether a paired rotational driver would also occur in that vicinity with a sensitivity of 100% (95%CI 81.5-100.0%) and specificity of 71.8% (95%CI 55.1-85.0%). The PPV and NPV was 62.1%, 95%CI 50.0-73.0% and 100% respectively.

**Figure 5A-D.**
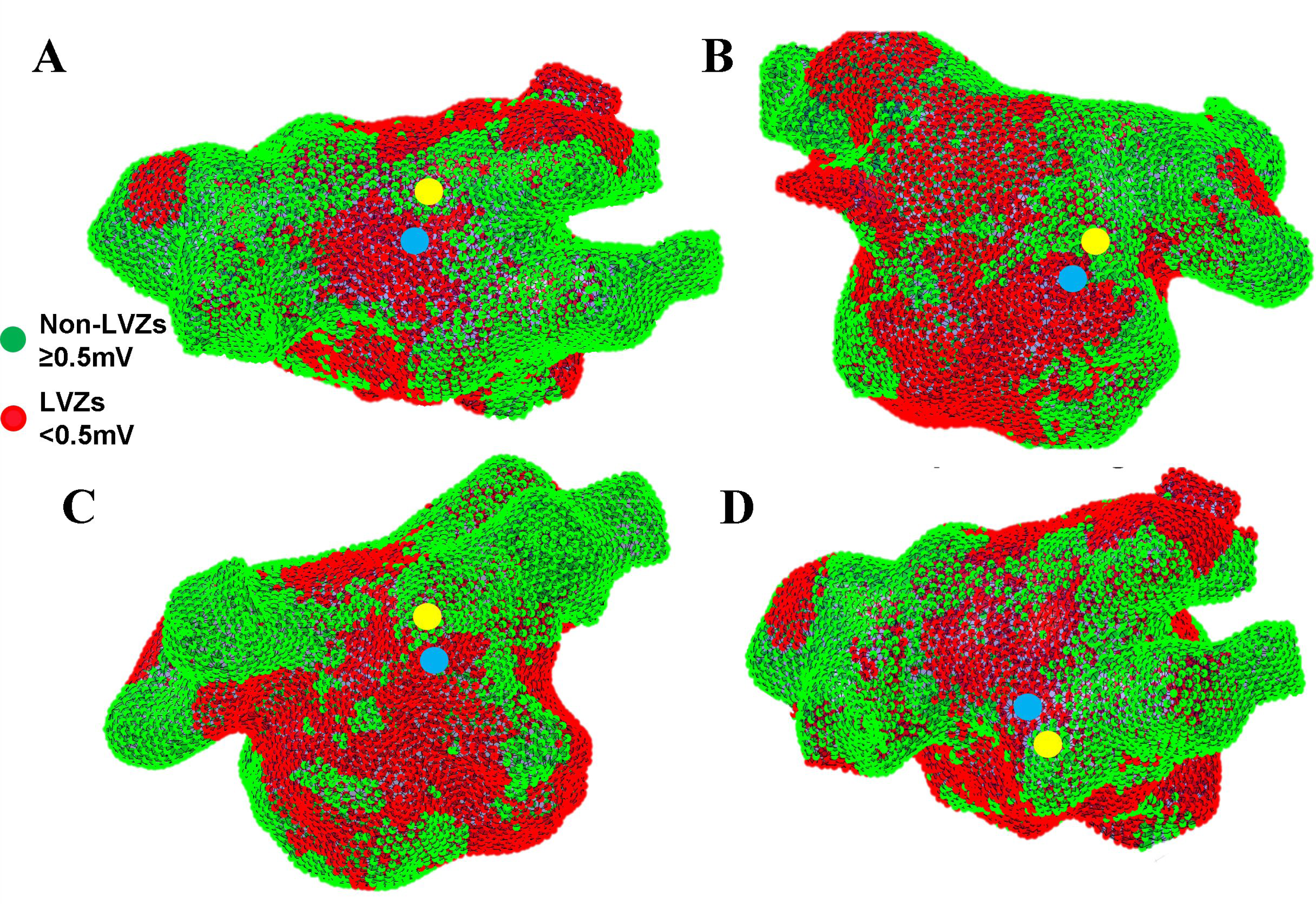
Demonstrates rotational (blue circle) and focal (yellow circle) drivers superimposed on a replica LA geometry and OV map created in EnsiteX. The rotational and focal drivers occur in close vicinity to each other whereby the rotational driver is mapped to a LVZ and the focal driver to the border of the site of scar in a nLVZ. Focal and rotational driver mapped to the ***A)*** roof on a LA map in a roof view, ***B)*** posterior wall on a LA map in a posterior-anterior view and ***C)*** anterior wall on a LA map in anterior-posterior view ***D)*** posterior wall on a LA map in a tilted posterior-anterior view.

Of the 48 rotational drivers, 35 (72.9%) were mapped to LVZ, and 32 of these (91.4%) were mapped to a close proximity to a focal driver. Rotational drivers were therefore more likely to be paired to a focal driver if the rotational driver was mapped to a site of LVZ (0/8, 0% nLVZ versus 32/35, 91.4% LVZ; p<0.001). Of the 8 rotational drivers mapped to nLVZ, none of which met our definition for pairing with focal drivers, the median geodesic distance to a focal driver was 33.2±9.2mm.

## DISCUSSION

### Main study findings

In this study, we have effectively mapped focal and rotational drivers in persistent AF that were spatially stable but with temporal periodicity. We also observed for the first time that although focal drivers often occur in isolation, rotational drivers have a strong spatial and temporal association with focal drivers, occurring mostly in close proximity to focal drivers and during or shortly after a burst of focal activations. Whilst focal drivers were shown to co-localize to nLVZs, rotational drivers co-localized to LVZs. Rotational drivers not paired to focal activations and occurring in nLVZs recurred less often (lower recurrence rate) and completed for fewer cycles at each occurrence (lower temporal stability). It has been shown drivers with a higher temporal stability and recurrence rate are more likely to result in AF termination (2,11) this potentially suggests that these are less mechanistic importance.

### Relationship between drivers and structural remodeling

In this study, we have shown that increasing AF duration is associated with a greater proportion of LVZs. This is consistent with the findings of other studies which have shown that AF duration correlates to the level of atrial structural remodeling (12,13). This study has shown that focal drivers in contrast to rotational drivers are more likely in those with less structural remodeling, and that rotational drivers may only occur in those with significant structural remodeling. Rotational drivers have in this study and previous studies shown to have a predilection to LVZs (1–4). A majority of rotational drivers in this study were mapped to LVZs. Rotational drivers mapped to LVZs demonstrated greater temporal stability and higher recurrence rate than rotational drivers mapped to nLVZs. This may suggest that rotational drivers mapped to LVZs play a greater mechanistic role in maintaining AF than those mapped to nLVZ.

It is unclear why this should be. One explanation might be that rotational drivers in nLVZ are functional and may represent true rotors, whereas those in LVZ are associated with structural remodeling and an anatomically defined circuit. These may therefore represent separate phenomena, and it appears that the latter is either more important or possibly is more easily eliminated with ablation.

In this study, a majority of drivers identified were focal in nature. Our previous work has shown that rotational drivers were more prevalent in persistent AF (1,2). The shorter AF duration in this study cohort in patients with healthy atria could account for the discrepancy seen. This again supports the hypothesis that rotational drivers are predominantly limited to areas of structural remodelling.

### Relationship between focal and rotational drivers

This is the first study to report a relationship between focal and rotational drivers in AF which is arguably an important evolution in our understanding of AF. Although focal drivers often occurred in isolation, rotational drivers were mostly found in close proximity to focal drivers and mostly occurred during or shortly after the focal activations. In fact, the presence of a LVZ near a focal driver was a strong predictor of whether it would be paired to a rotational driver.

The mechanisms underlying what is described as rotational drivers remain unclear. Rotational drivers mapped to nLVZs are not paired to focal drivers, are less temporally stable and recur less. This may suggest they are more likely to be sustained by functional re-entry and may be less mechanistically important. However, the observation that rotational drivers mapped to LVZs mostly occur in close proximity to focal drivers arguably suggests anatomical re-entry, whereby the focal driver triggers re-entry. The temporal relationship, with the focal driver preceding the rotational driver in a majority of cases also supports this conclusion.

### Implications for ablation strategy

Others have used a voltage-guided substrate modification technique which adapts the AF ablation strategy to the underlying bipolar voltage map (14–17). In these studies, patients with LVZs will undergo further ablation to homogenize scar in addition to PVI whilst those with healthy atria have no further ablation beyond PVI. Utilizing this approach may eliminate many of the rotational drivers which depend on structural remodeling. However, it may not be sensitive or specific for driver sites and may create further scar and predispose to ATs. It may be possible to refine this approach by targeting LVZ associated with rapid activity in AF (16), although there are conflicting data regarding the usefulness of electrogram characteristics or CL as a surrogate marker for sites driving AF (1,5,6). Another possibility might be to examine conduction velocity dynamic in LVZs in SR to determine whether they are likely to support rotational activity in AF (3).

A major limitation to a strategy targeting LVZs is that it will not address the focal drivers observed in this study. Focal activations may represent different phenomena such as micro-reentry or intramural re-entry, or perhaps given the lack of association with LVZs they may represent sites automaticity and co-locate to sites of ganglionated plexi innervation. Regardless of the underlying mechanism, focal drivers may be legitimate ablation targets in AF. It is therefore likely that a strategy targeting either focal or rotational drivers alone (either directly or through surrogates) will miss mechanistically important targets.

A better understanding of the mechanisms of localized drivers will ensure the ablation strategy is adapted to the stage of AF. In patients with healthy atria, focal drivers were predominantly identified and therefore in this context an ablation strategy prioritising focal drivers would seem ideal. However, in scarred atria rotational drivers were identified thereby suggesting that rotational drivers should be targeted in addition to focal drivers. Further to this, the higher temporal stability and recurrence rate of rotational drivers paired to focal drivers potentially indicated they are mechanistically more important in AF and should be prioritised with ablation in a hierarchal approach.

These patient specific mechanisms may explain difficulty treating AF with current standardised approaches. Further randomized controlled trials are needed to evaluate driver-guided ablation in different clinical protocols.

### Limitations

The focus of this study was to evaluate the relationship between focal and rotational drivers to underlying scar and to each other. In this study, the drivers were not ablated and therefore their mechanistic importance in AF with regards to electrophysiological endpoints has not been established. However, we have previously shown that a majority of drivers identified using this definition for a focal and rotational driver result in an electrophysiological response on ablation and thereby indicating an important mechanistic role in AF (2,11,18).

## CONCLUSIONS

These data demonstrate that intermittent localized focal and rotational drivers occur in patients with persistent AF post-PVI. Although focal drivers seem unaffected by the presence or absence of structural remodeling, rotational drivers were largely dependent on the underlying substrate. Rotational drivers are often paired to focal drivers and occur when focal drivers are in close proximity to LVZs. The higher temporal stability and recurrence rate of rotational drivers mapped to LVZs and, when paired with a focal driver compared to rotational drivers that are not is suggestive that these are mechanistically more important in AF and should be targeted as a priority allowing a hierarchical approach to driver ablation. The relationship between rotational and focal drivers also emphasises the importance to target both during localized driver ablation of AF. These novel mechanistic observations require further scrutiny using different mapping and imaging modalities but outline a plausible model for patient specific mechanisms maintaining AF.

## Data Availability

The data underlying this article will be shared on reasonable request to the corresponding author.

## Notes

### Author Declarations

Patients provided informed consent for their study involvement which was approved by the UK National Research Ethics Service (22/PR/0961).

